# Low-Cost Differential Pressure Spirometry for Emergency Ventilator Tidal Volume Sensing

**DOI:** 10.1101/2020.09.25.20198069

**Authors:** Jordan L. Edmunds, Mauricio J. Bustamante, David K. Piech, Jonathan S. Schor, Samuel J. Raymond, David B. Camarillo, Michel M. Maharbiz

## Abstract

COVID-19 has become a significant burden on the healthcare systems in the United States and around the world, with many patients requiring invasive mechanical ventilation (IMV) to survive. Close monitoring of patients is critical, with total volume per breath (tidal volume) being one of the most important data points. However, ventilators are complex and expensive devices, typically in the range of tens of thousands of US dollars, and are challenging to manufacture, typically requiring months. Solutions which could augment the ventilator supply rapidly and at low cost in the United States and elsewhere would be valuable. In this paper, we present a standalone tidal volume measurement system consisting of a D-Lite spirometer, pressure sensor, microcontroller, and tubing with a cost of parts less than $50 USD. We also provide a model to predict the error in tidal volume measurements based on the pressure sensor used and the flow during ventilation. We validate this system and show that the tidal volume accuracy for flows above 10 L*/*min was within 10%. We envision this system being used to increase the ventilator supply in resource-constrained settings.

## 1 Introduction

As of August 25, 2020, there have been over 23 million cases and 815,000 deaths from the COVID-19 disease caused by the SARS-CoV-2 virus[1], with hospitals across the world shouldering the burden for treatment in most severe cases. In the United States, of those who are admitted to the hospital for COVID-19, over 20% are subsequently admitted to the intensive care unit (ICU), although this number is decreasing as we learn how to better treat this disease. The majority of these patients (69%) require invasive mechanical ventilation (IMV) for a mean duration of 10 days [2].

Monitoring of mechanical ventilation is critically important to ensure patients are receiving enough air. Typically, this is done by delivering a set volume of air to the patient in a given breath (tidal volume) based on their body weight [3]. Insufficient volume can lead to hypoventilation and CO2 buildup, or atelectasis (lung collapse). From the onset of the pandemic, there were concerns about equipment shortages, including intensive care unit (ICU) ventilators [4, 5]. This led to some companies re-purposing facilities to participate in ventilator production [4], the U.S. president invoking the Defense Production act [6], as well as a number of emergency ventilator design efforts to bypass supply chain issues [7, 8, 9, 10]. The shortage was so severe governmental agencies released emergency guidelines, including an FDA guidance allowing clinicians to repurpose BiPAP (bilevel positive airway pressure) machines commonly used as a treatment for sleep apnea, anesthesia machines, and simpler ventilators for emergency use if needed [11, 12].

These events have made clear the importance of developing a low-cost, accurate, and easily deployed system that can supplement the existing IMV supply chain in future catastrophes. Most existing efforts to this end are focused on ventilators which are very cheap and but only provide pressure measurements [7, 8], or on designs that rely on external and expensive flow meters [8]. In this work, we present a middle ground to these two approaches: a guide to designing a cheap and accurate flow sensor for measurement of tidal volume, based around an easy-to-manufacture plastic spirometer which has been around for several decades. The advent of accessible microcontrollers and the rapidly falling cost of silicon electronics enable a low-cost, high-accuracy tidal volume measurement system to be built with interchangable parts which could supplement the functionality of existing minimal-feature ventilators or alternative respiratory devices, such as BiPAP machines. We envision such a device could be crucial in future pandemics and may also serve an important role in current low-resource settings.

## 2 Methods

Our tidal volume meter consists of three main components: (a) a passive spirometer, (b) a differential pressure sensor, (c) a computational unit. Figure 1 shows how these components interact with each other and a larger system.

**Figure 1:**
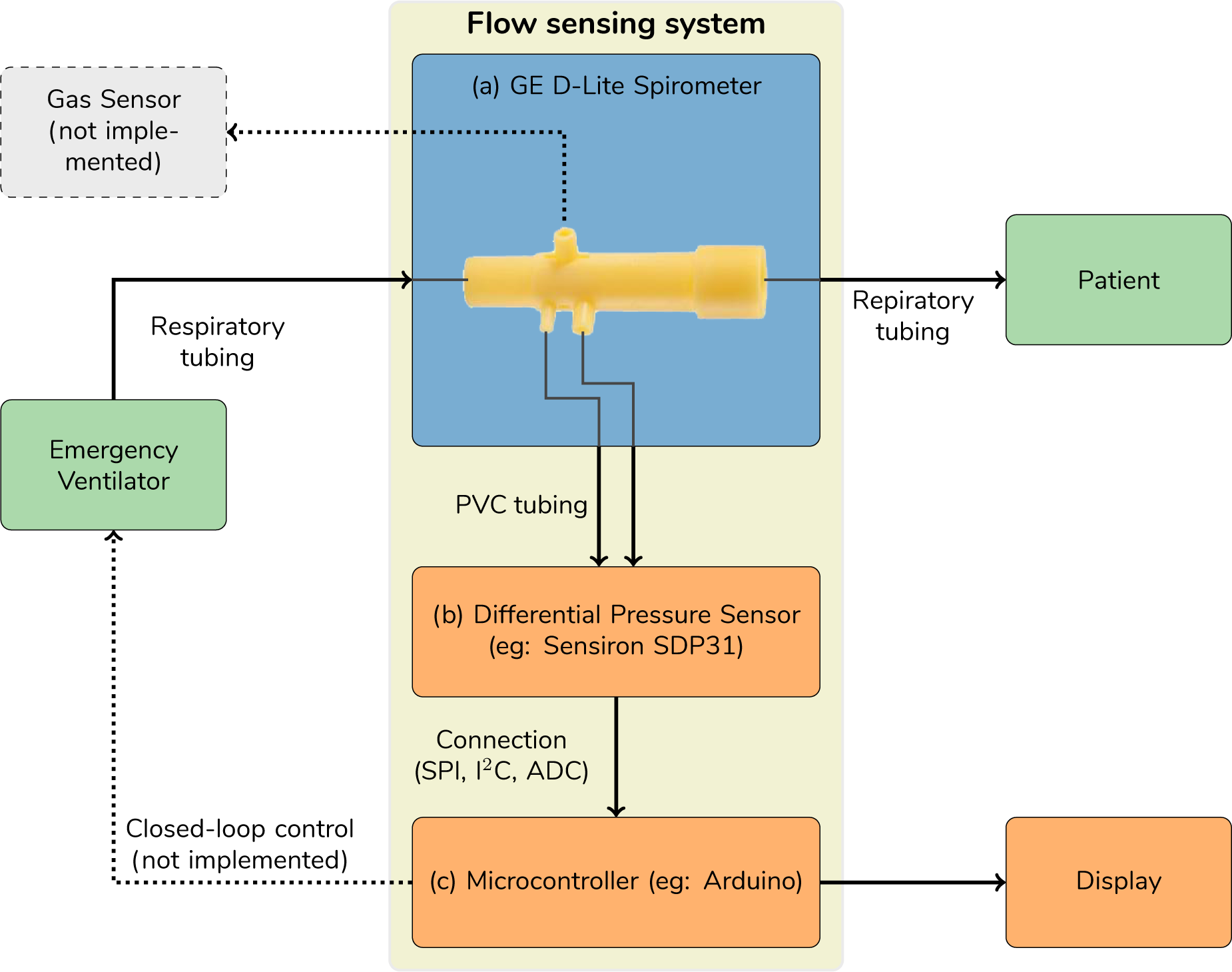
Overview of flow sensing system and integration in a clinical setting. The D-Lite spirometer fits into sections of standard 22 mm ventilator tubing (with an adapter at the smaller end). The dynamic pressure drop is sensed across the two oppositely-directed terminals by a pressure sensor (e.g. the Sensiron SDP31), and this information is sent to a central processing unit (e.g. an Arduino). This information can then be displayed or used in closed-loop feedback.

### 2.1 Design Considerations

For the passive spirometer, we selected the General Electric D-Lite ++ spirometer [13, 14], which transduces flow velocity into a pressure difference. Although this part is available online, we developed a similar model that can be 3D printed (see Suplementary information). The device has two Pitot tubes, one upstream of a flow constriction and facing upstream, and the other downstream of the flow constriction and facing downstream, and a third port for gas sensing.

The dynamic pressure between between both Pitot tubes depends on the flow by:

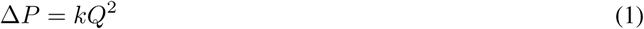

where Δ*P* is the pressure difference in cmH_2_O, and *Q* is the flow in L*/* min. A proportionally constant *k* in cmH_2_O*/*(L*/*min)^2^ depends on air density, the spirometer channel geometry, and the Reynolds number [13]. Since the pressure difference also changes sign for reverse flow, there is a one-to-one correspondence between flow and pressure. However, due to the quadratic nature of the relationship, measurements at very low flows are challenging. Most regulations require reporting of tidal volume to a given relative accuracy, such as 10 % [12]. In order to achieve this, a sufficiently accurate pressure sensor must be selected, and the error should be quantified. The error in volume measurements can be found integrating equation (1) over time, and assuming some absolute pressure accuracy. We show this accuracy in Figure 2 as a function of sensor pressure accuracy and flow.

**Figure 2:**
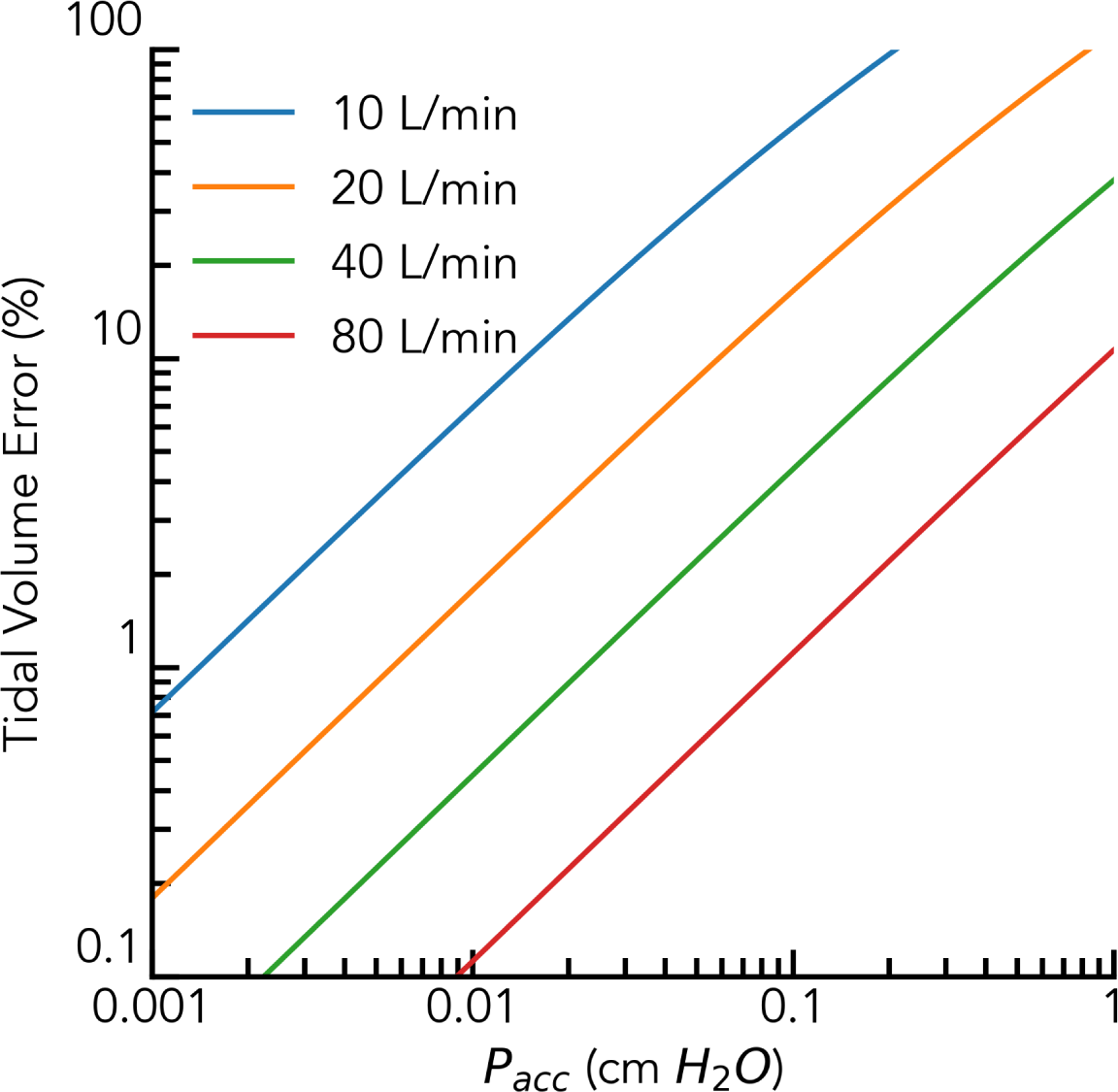
Tidal volume error vs. pressure sensor absolute accuracy at varying flows. Each curve corresponds to a different instantaneous flow (in which case the results are exact), or average flow over a sensing period, in which case they are approximate. The Sensiron SDP31 has an absolute accuracy of 0.001 cmH_2_O, while the Honeywell SSCMRRN060MDSA5 has an accuracy of 0.01 cmH_2_O. Note that the error increases with worsening accuracy and lower flow

Among electronic pressure sensors, the key features involve communication protocol, pressure range, wide commercial availability, and accuracy, as a percentage of full scale. We selected digital pressure sensors to minimize part count and the need for a PCB, and settled on the Sensiron SDP31 (0.001 cmH_2_O absolute accuracy) and a less accurate Honeywell Honewell SSCMRRN060MDSA5 (−6 kPa to 6 kPa, 0.25% of full scale accuracy).

A microcontroller or similar computational unit is needed to compute flow and integrate it over time for tidal volume. Many emergency ventilators already have a comparable device for control and sensing [9, 10]. We chose an Arduino for its well-supported software ecosystem and wide availability, and these can be purchased off-the-shelf for as low as $5. The Arduino runs a finite state machine (see Sumplementary Materical) to read the pressure, convert it into flow, detect breath onset with a simple threshold, and compute tidal volume, with a sampling and computational frequency of 167 Hz. This is then reported to a computer for display or further analysis. The spirometer was calibrated by comparing pressure measured with both sensors against flow, where the flow was measured with a gold standard Alicat M-Series 1000 SLMP mass flow meter.

### 2.2 Pressure Sensor and Ventilator Characterization

We connected the Honewell differential pressure sensor to an Arduino using SPI and measured data at 166 Hz. To ensure the Honeywell sensor was operating as expected, we measured its pressure reading against that of a water-column manometer in the range of 0-4 cmH_2_O (Supplementary Figure S1) by varying the flow through the spirometer. This information was used to correct an offset of −0.02 cmH_2_O. Having validated the sensor was behaving as expected, we characterized the pressure of the D-Lite spirometer as a function of flow using both the Honeywell sensor (0.01 cmH_2_O absolute accuracy) and a more accurate sensor (Sensiron SDP31, (0.001 cmH_2_O water absolute accuracy). We then offset-corrected data from the Honeywell and the Sensiron and fitted the resulting flow vs. pressure to a quadratic function (Figure 3). We additionally corrected the Sensiron data for the pressure drop from the measurement tubes (1/16in ID, 1 meter length) using a company-provided datasheet with correction factors [15].

**Figure 3:**
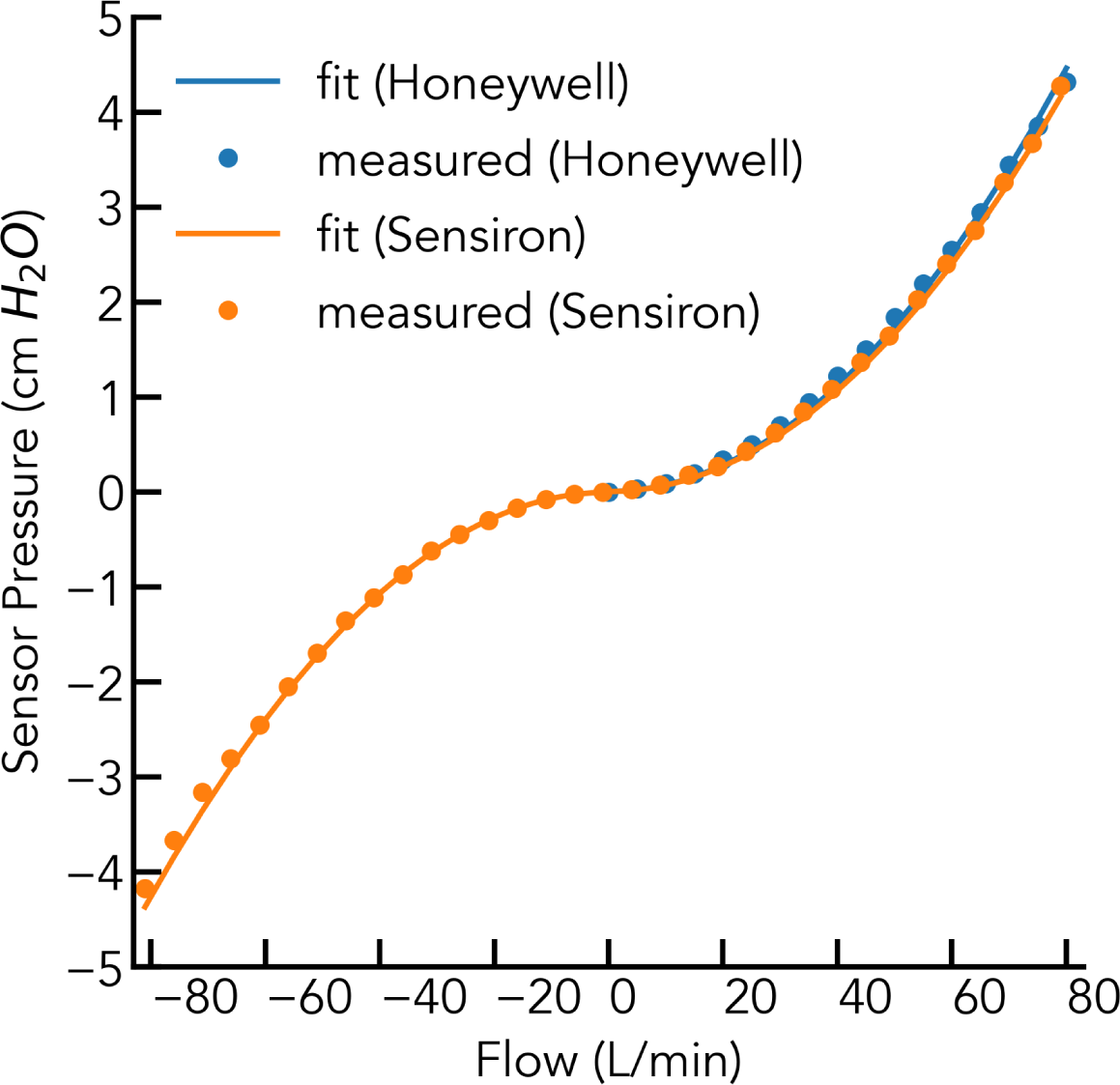
Pressure vs. flow for the D-Lite spirometer as measured by Alicat mass flow meter. Honeywell data is corrected for zero offset prior to fitting to quadratic. Uncertainty in flow is <3 L*/*min at all flows, and uncertainty in pressure is <0.01 cmH_2_)

We investigated transient behaviour for typical ventilator breaths by connecting the spirometer to a Bird Mark 7 pressure controlled ventilator and a rubber test lung (Figure 5) with the Honeywell sensor. Similarly, we measured the transient behaviour for a constant flow emergency ventilator (an early prototype of the Stanford Rapid Response ventilator) [16] and a Dua-Adult Training and Test Lung Model 1600 (Michigan Instruments) (Figure 4). The device interrupts pressurized flow, with a constant-flow valve, which drops a regulated 50 psi of pressure across it. The spirometer was connected in several different locations: the inspiratory limb, expiratory limb, and near the test lung such that it recorded inspiration only, expiration only, and both inspiration and expiration for each breath. We tested nominal flows (set by a needle valve and determined by a rotameter) of 10, 30, and 50 L*/*min as well as lung compliances of 10, 30, and 150 mL*/*cmH_2_0, with at least 138 breaths in each condition. Each breath lasted 4 seconds with a 1:3 inspiration to expiration ratio (I:E).

**Figure 4:**
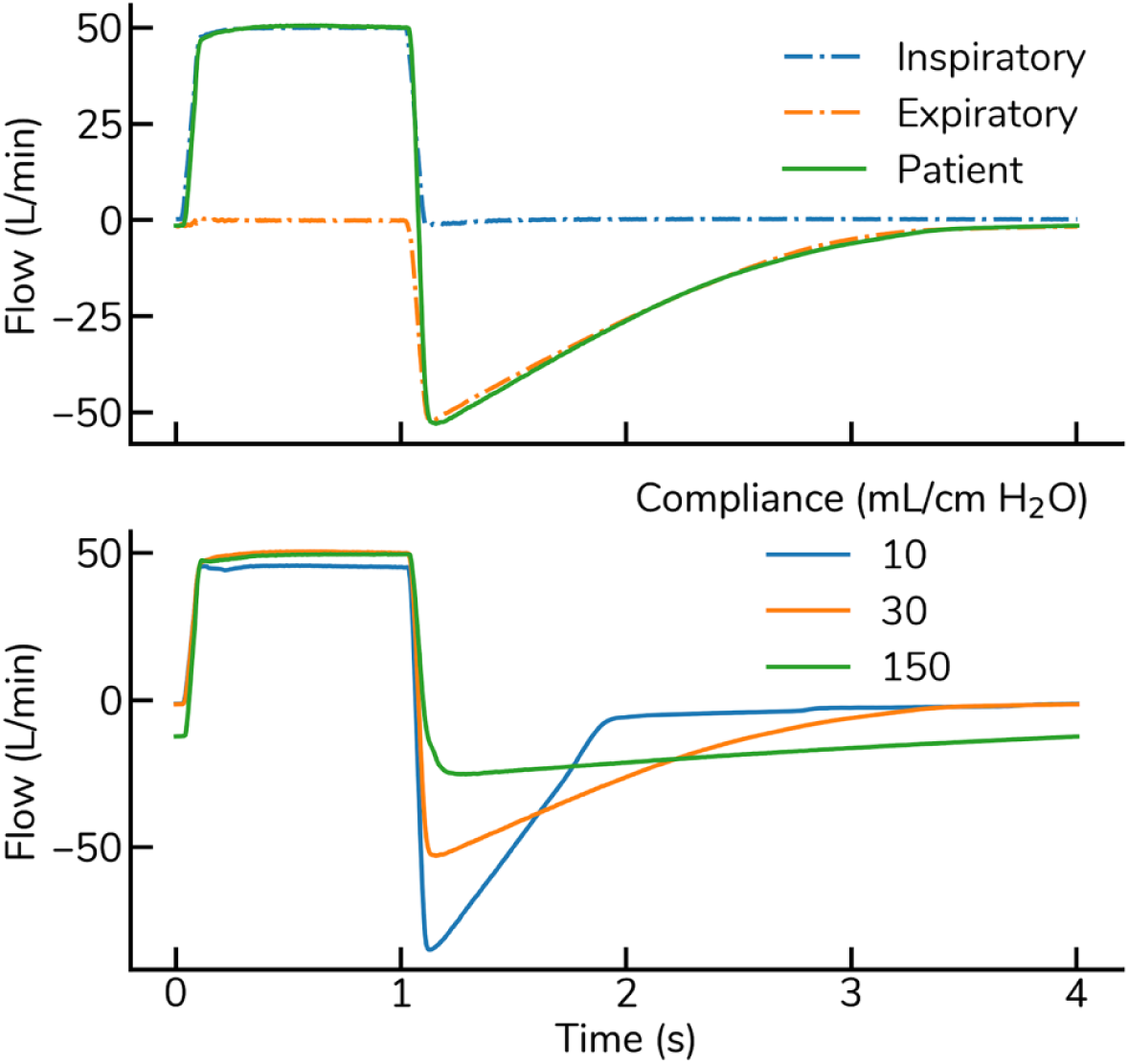
Flow vs. time over a single 4-second breath cycle for the Stanford Rapid Response ventilator. Data was obtained from 138 breaths and averaged into a single plot (top). Test lung compliance was varied at a fixed flow, and the resultant flow profiles are plotted versus time (bottom).

**Figure 5:**
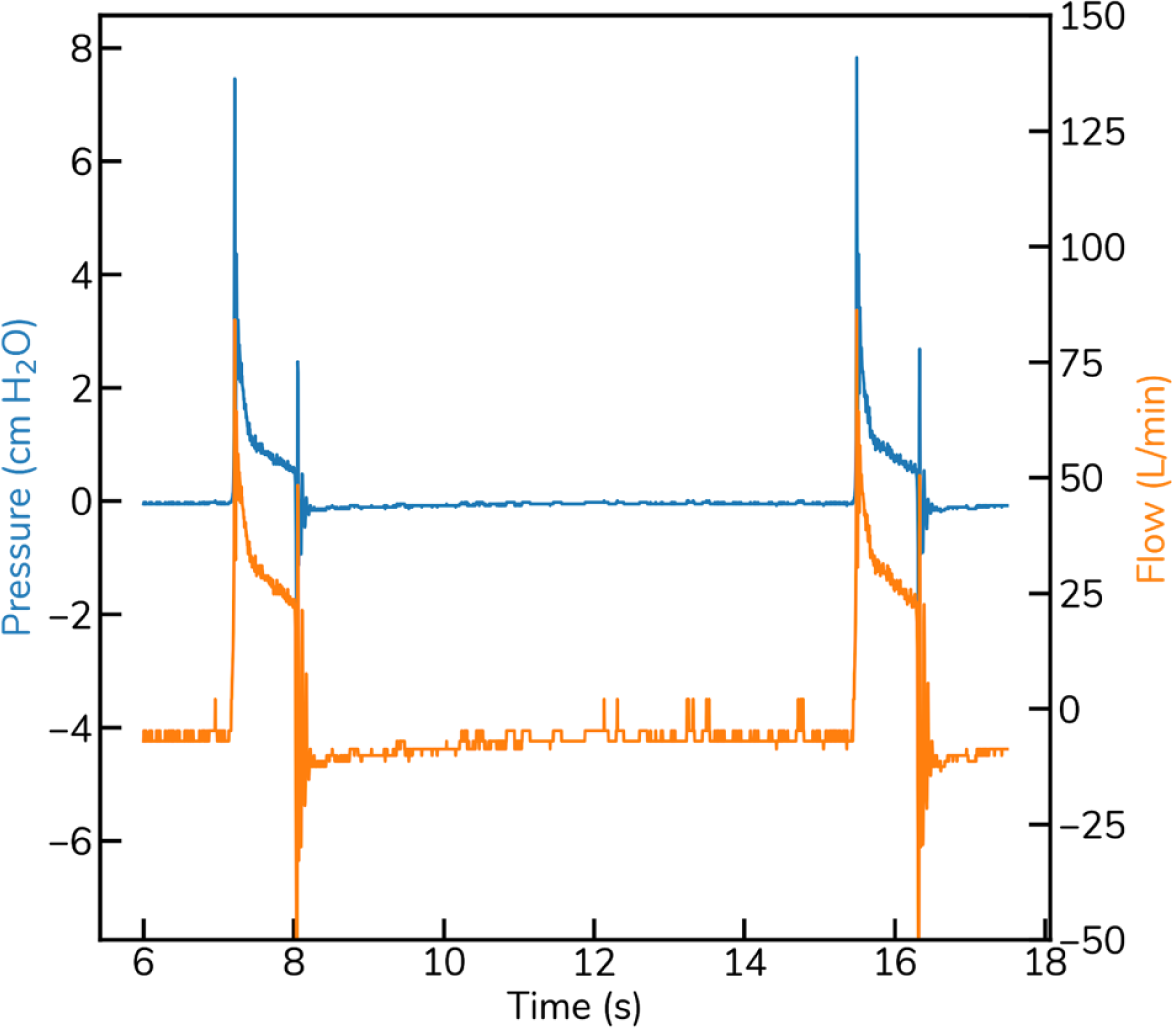
Pressure and derived flow over two sample breaths given by a Bird Mark 7 to a simple test lung, as recorded by the Honeywell differential pressure sensor.

## 3 Results

We found the proportionality constant k to be 6.97*10-4 6.97 *×* 10^*−*4^ cmH_2_O*/*(L*/*min)^2^ for the Honeywell with a standard error of the regression of 0.06 cmH_2_O, and 6.43 *×* 10^*−*4^cmH_2_O*/*(L*/*min)^2^ for the Sensiron with a standard error of the regression of 0.03 cmH_2_O prior to pressure drop correction. After this correction, the proportionality constant increased to 6.7 *×* 10^*−*4^ cmH_2_O*/*(L*/*min)^2^.

Once the proportionality constant was known, we used this to measure flow from the Stanford Rapid-Response ventilator at various flows (Figure 4). The measurements were very repeatable, with maximum flow standard deviations of 0.5% and and min/max differences of 2% of the expected volume (see Figure S2). To ensure our thresholding method was accurate, we verified it agreed with offline numerical integration (Figure S3). We also characterized an older model pressure-controlled ventilator, the Bird Mark 7, and show a typical breath curve in both pressure and volume (Figure 5). Finally, we characterized the accuracy of our system at measuring tidal volume using a 500mL syringe, varying the breath time and volume (Figure 6). The accuracy of these measurements is compared to our accuracy model (Figure 7).

**Figure 6:**
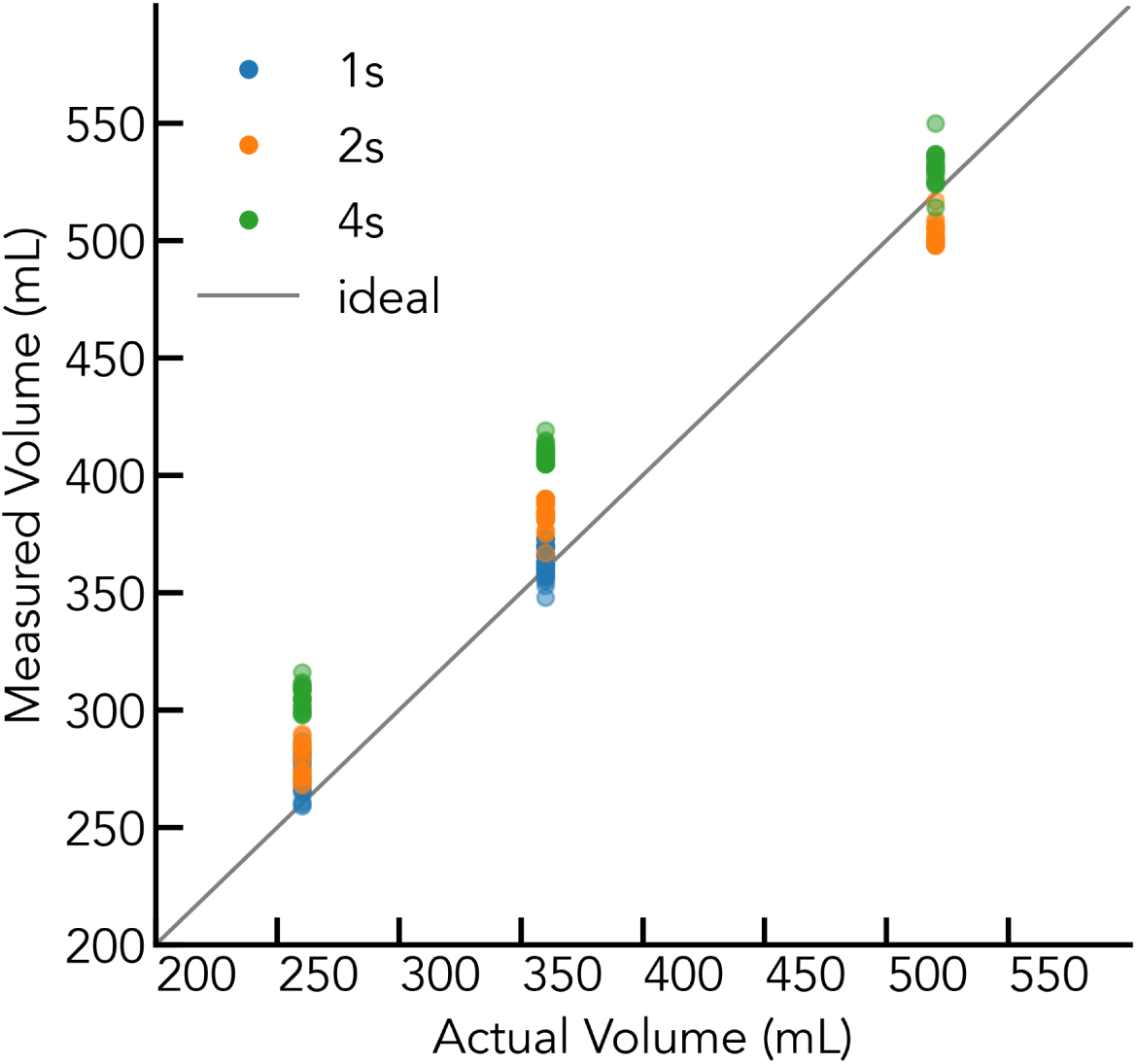
Measured volume from syringe experiments using 1s, 2s, and 4s flow durations. Error increases at lower flows, with a maximum error of MAX at the lowest volume/flow rate (3.75L/min), much lower than typical flow for an adult ventilator. Syringe plunges were performed visually, and the error increases with decreasing flow. Expected maximum error from device alone is 6% at the lowest flow, and the mean of the lowest-flow measurements are 10% away from the mean of the highest-flow measurements in the same condition.

**Figure 7:**
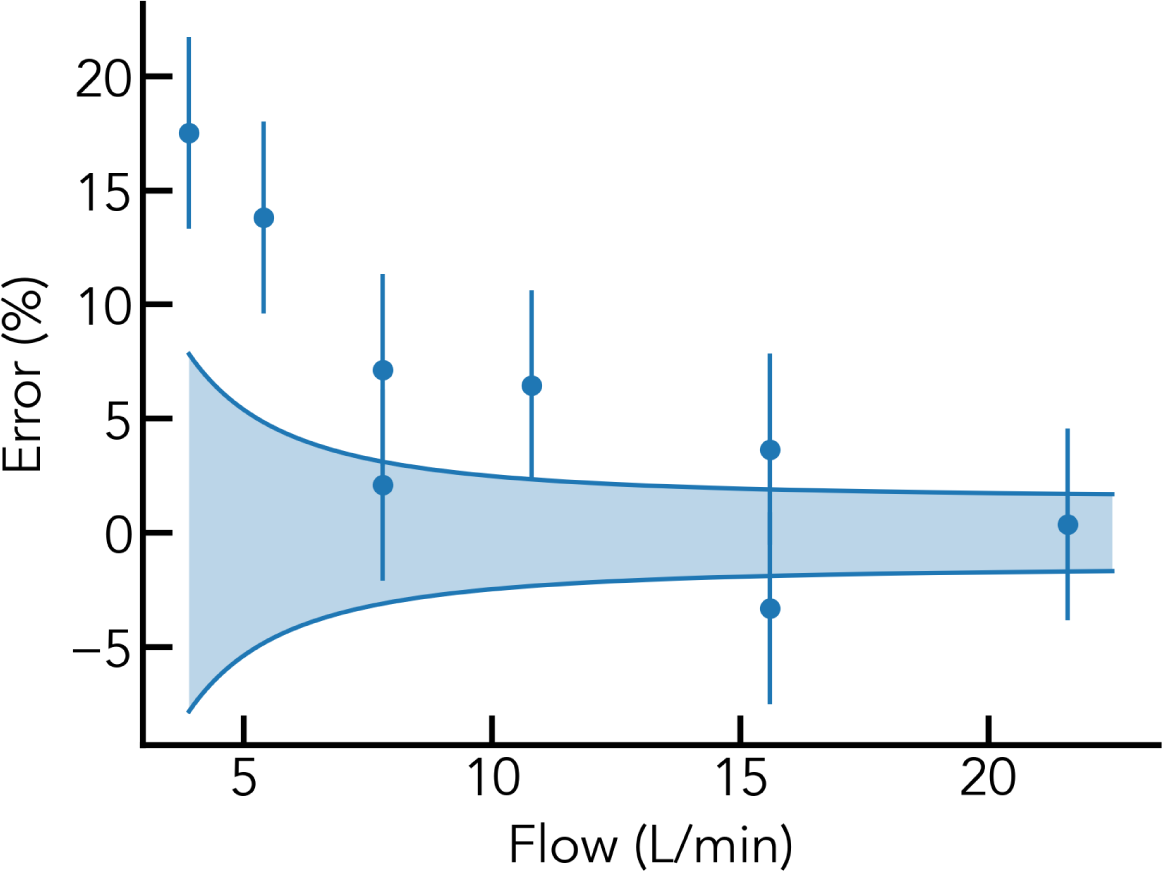
Error vs. Measured flow taking the mean of inhaled and exhaled breaths (10 trials each, 20 total measurements per condition)

## 4 Discussion

We designed a low-cost differential-pressure based system to measure inhaled and exhaled flow based around the D-Lite differential pressure spirometer, which consists of a microcontroller, differential pressure sensor, D-Lite spirometer, and connecting wires and tubing. We used this system to characterize the D-Lite spirometer, and report the measured differential pressure vs. flow for two different pressure sensors. We measured the proportionality constant *k* to be between 6.43× 10^*−*4^ and 6.97 × 10^*−*4^ cmH_2_O*/*(L*/*min)^2^. We also developed a model to estimate the error in converting the differential pressure measurement to flow, depending on the pressure sensor used, and used this to predict the relative volume error vs. flow (Figure 7). The accuracy of our measurements of tidal volume exceeded the expected error of our sensor model, indicating there are unaccounted-for sources of experimental error which are not due to the sensor alone. The main source is the error in the proportionality constant, as any error in that constant is (for small errors) linearly proportional to the error of a volume measurement. This level of accuracy is sufficient for adult breath measurement clinical use cases where flows tend to be larger (>10L/min). With a cost of components of <$50, such a system could be a practical add-on to otherwise simple mechanical ventilators in emergency or low-resource settings.

If high accuracy is needed at both low and high (relative to full-scale) flows, more than one pressure sensor may be used, one with a small full-scale range to ensure accuracy at low flows, and the other with a large full-scale to capture larger flows. This, however, increases the cost of the system, as the pressure sensor is the most expensive component.

As a proof of concept, we used the D-Lite spirometer and SDP31 differential pressure sensor to characterize the operation of the Stanford emergency response ventilator [16] (a flow-controlled ventilator), and the Bird Mark 7. As can be seen in Figure 6, at flows near zero the flow discretization noise becomes very large (the derivative of the square root function becomes very large near zero). Even a zero offset of a few least significant bits (a very good pressure sensor) can correspond to a flow offset large enough to significantly affect the measured tidal volume.

We also demonstrated that our simple finite state machine based thresholding for calculating tidal volume on the Stanford rapid response ventilator was nearly identical to the performance of offline integration, with deviations of at most 2% from the integrated value, with a standard deviation of 0.5%. This was true across settings for typical flows (10, 20, and 50 L*/*min) and lung compliances (10 - 150 mL*/*cmH_2_O per lung) for a typical lung resistor (Rp20). As long as the sensor is sufficiently accurate so that the zero offset is below the minimum expected measurable flow, this should yield accurate tidal volumes for flow-controlled ventilators.

Taken together, these results demonstrate that a simple, low-cost system can be used to accurately determine tidal volume independently of the ventilator used. This could supplement the functionality of existing ventilators or alternative respiratory devices, and may help increase the ventilator supply in future pandemics or in low-resource settings.

## 5 Conclusion

We designed a differential pressure based flow measurement system around the D-Lite spirometer. This system was shown to be accurate to within 15% for measurement of tidal volumes for all conditions tested (4L/min - 20L/min), and was used to characterize the Stanford rapid response ventilator. The data and methodology we present here could be used to design simple, low-cost flow measurement systems.

## Data Availability

All raw data is available on our group's google drive. Code used for online data processing can be found on github. Both links are available below.

https://drive.google.com/drive/u/0/folders/1wQxoaZd3DSQLKDs2eLh8qEjYdGccdzsO

https://github.com/edmundsj/TidalVolumeArduino

## 6 Acknowledgements

We would like to thank the UCSF COVID-19 Response Fund for supporting this work, as well as the Chan-Zuckerberg Biohub and Hertz Foundation for supporting this work. We would also like to thank members of the Stanford Linear Accelerator (SLAC), especially Professor Aaron Roodman and his team for useful conversations, in addition to members of 219 Design.

## Suplementary Material

### Code and designs

Code for the Arduino and data processing is available at https://github.com/edmundsj/TidalVolumeArduino. Raw data is available at https://drive.google.com/drive/u/0/folders/1wQxoaZd3DSQLKDs2eLh8qEjYdGccdzsO.

Open source 3D printed designs for spirometry can be found at https://3dprint.nih.gov/discover/3dpx-013632.

**Figure S1:**
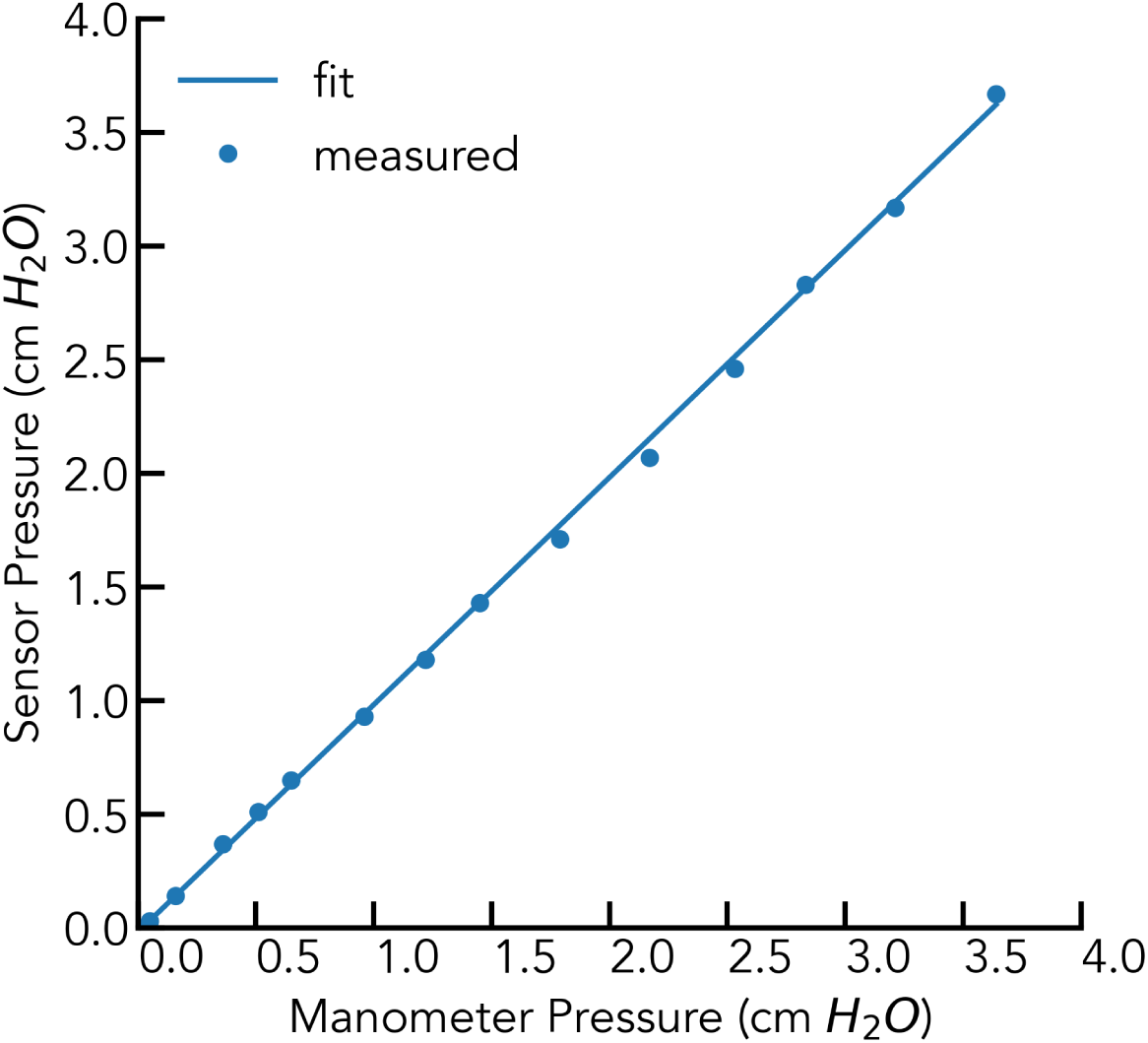
Measured pressure sensor data vs. data taken from water column manometer. Fit line is *y* = 1.000*x*− 0.0185, where *x* is the measured manometer pressure and *y* is the sensor pressure. Pictures of the water column manometer were taken using a stationary iPhone 11 at 2.7x optical zoom, and analyzed using ImageJ to find the height of the water.

**Figure S2:**
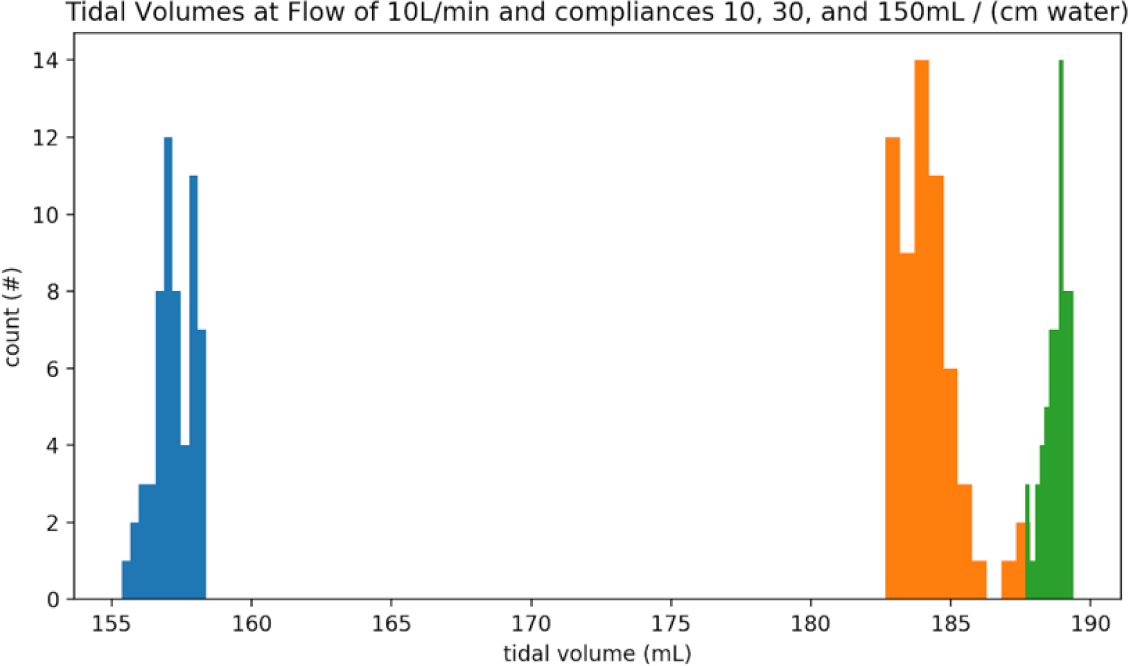
Histogram of measured tidal volumes of Stanford rapid-response ventilator at compliances of 10, 30, and 150 mL*/*cmH_2_O at the lowest flow characterized (10 L*/*min). The intra-compliance variability was <2% min/max with standard deviation <0.5%

**Table S1:** Bill of Materials

| Part Name | Source | Cost | Quantity | Total Cost |
| --- | --- | --- | --- | --- |
| Arduino Due * | Amazon | \$22 | 1 | \$22 (\$5) |
| Arduino Nano | Amazon | \$5 | 1 | \$5 |
| D-Lite Spirometer | 4MDMedical | \$8ea | 1 | \$8 |
| Pressure Sensor (SDP31) | Digikey | \$31 | 1 | \$31 |
| Medical-grade tubing | US Plastics | \$1/ft | 1ft | \$1 |
| **Total** | | | | **\$62 (\$45)** |

\* The Arduino Due can be substituted for the Arduino Nano at a cost of \$5.

**Figure S3:**
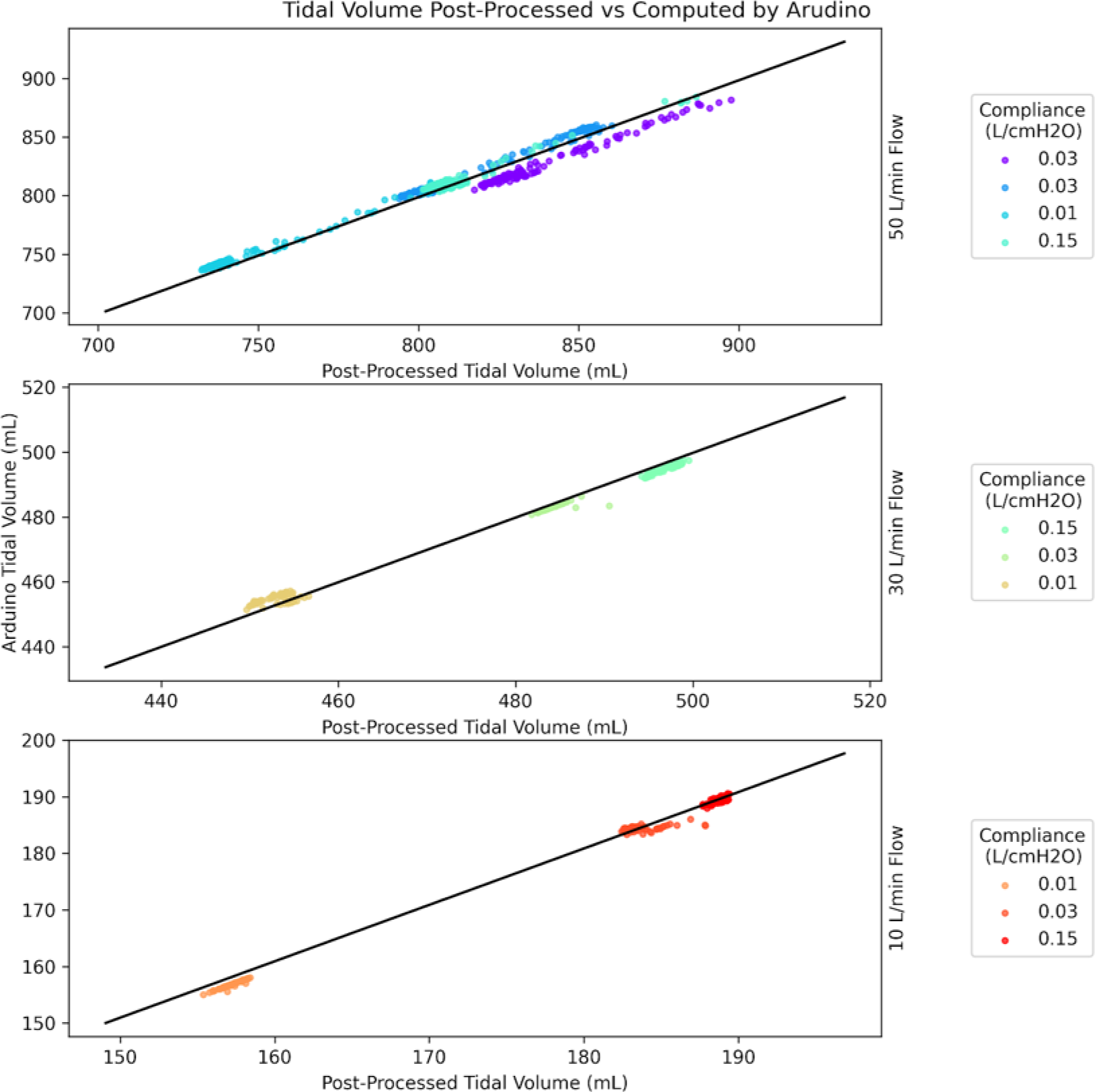
Tidal volumes computed by offline numeric integration vs. tidal volumes computed using online thresholding algorithm. Graphs are for three different flows (50L/min, 30L/min, and 10L/min).

## Notes

### Competing Interest Statement

The authors have declared no competing interest.

### Funding Statement

An ongoing grant from the Chan Zuckerberg Biohub funded the graduate students involved in the work. Money used to purchase most of the supplies used was from the UCSF COVID-19 Response Fund. One author is receiving ongoing support from the Hertz Foundation. No external services were given by third parties or were involved in the study design except as stated by the Acknowledgements.

### Author Declarations

As this paper describes the design and analysis of a medical devices that was not tested on humans no IRB approval was requested.

